# Causal effects of breast cancer risk factors across hormone receptor breast cancer subtypes: A two-sample Mendelian randomization study

**DOI:** 10.1101/2024.09.02.24312928

**Authors:** Renée MG Verdiesen, Mehrnoosh Shokouhi, Stephen Burgess, Sander Canisius, Jenny Chang-Claude, Stig E Bojesen, Marjanka K Schmidt

## Abstract

**Background:** It is unclear if established breast cancer risk factors exert similar causal effects across hormone receptor breast cancer subtypes. We estimated and compared causal estimates of height, body mass index (BMI), type 2 diabetes, age at menarche, age at menopause, breast density, alcohol consumption, regular smoking, and physical activity across these subtypes.

**Methods:** We used a two-sample Mendelian randomization approach and selected genetic instrumental variables from large-scale risk factor GWAS. Publicly available summary-level data for the following subtypes were included: luminal A-like; luminal B/HER2-negative-like; luminal B-like; HER2-enriched-like; triple negative. We employed multiple methods to evaluate the strength of causal evidence for each risk factor-subtype association.

**Results:** Collectively, our analyses indicated that increased height and decreased BMI are probable causal risk factors for all five subtypes. For the other risk factors, the strength of evidence for causal effects differed across subtypes. Heterogeneity in the magnitude of causal effect estimates for age at menopause and breast density was explained by null findings for triple negative tumours. Regular smoking was the sole risk factor for which there was no evidence for a causal effect on any subtype.

**Conclusions:** This study suggests that established breast cancer risk factors differ across hormone receptor subtypes.

## Introduction

Previous case-control and cohort studies provide some evidence for breast cancer subtype-specific risk factors, including reproductive factors ^1–3^, body mass index (BMI)^2,4,5^, and alcohol consumption ^2,6,7^, although reported associations are inconsistent across studies. A likely explanation for this inconsistency is the relatively small number of cases for rarer subtypes like HER2-enriched and triple negative breast cancer. In general, breast cancer studies that collected both risk factor data and detailed pathology data for a large number of women are limited. A second challenge is that results from observational studies are often subject to bias due to (residual) confounding, measurement error, and reverse causation ^8^. As a result, it remains unclear whether previous results reflect causal associations with breast cancer subtypes.

Mendelian randomization (MR) is a specific type of instrumental variable (IV) analysis that minimizes the risk of these biases through the use of germline genetic variants, provided that certain assumptions are valid ^8^. Previous MR studies on breast cancer risk supported causal associations for height ^9^, BMI ^10^, age at menarche ^11^, age at menopause ^12^, breast density ^13^ and physical activity ^14^, but not for type 2 diabetes (T2D)^15^, alcohol consumption and smoking ^16^. Because of the heterogeneity of breast cancer, also within estrogen receptor (ER)-defined subtypes, it is essential to assess causality of these associations with hormone receptor subtypes. Thus far, only a handful of MR analyses included data on these biologically more homogenous subtypes ^17–22^. However, most of these studies only partially assessed if all MR assumptions were met, and thus a comprehensive evaluation of causal evidence for subtype-specific risk factors is currently lacking.

Therefore, the aim of this study was to estimate and compare causal effects of nine established risk factors, including anthropometric, reproductive, and behavioural exposures, across five hormone receptor breast cancer subtypes using a two-sample MR design.

## Methods

### Study design: two-sample MR

We used a two-sample MR study design and summary-level data for both the risk factors and outcomes of interest. Specifically, we extracted summary statistics for single nucleotide polymorphism (SNP)-risk factor and SNP-breast cancer subtype associations from different genome-wide associations studies (GWAS), which are described into more detail below. All included GWAS conducted comprehensive quality control of the genetic data. To yield valid causal estimates, selected genetic IVs should meet the following three MR assumptions: (1) IVs are robustly associated with the risk factors of interest (relevance assumption); (2) IVs are not associated with confounders of the studied associations (independence assumption); and (3) IVs do not affect the risk of breast cancer subtypes through mechanisms that do not involve the risk factors of interest (exclusion restriction assumption)^8^. An important additional assumption of two-sample MR is that the study participants included in both samples are from similar underlying populations (homogeneity assumption)^23^. We performed extensive secondary analyses to assess if these assumptions were reasonable.

### Data sources for SNP-risk factor associations

In 2020, Cancer Research UK published a list of established breast cancer risk factors, based on scientific evidence up to that moment ^24^. From this list we selected the following nine breast cancer risk factors for our MR analyses: height, BMI, T2D, age at menarche, age at menopause, percent breast density, alcohol consumption, regular smoking, and overall physical activity. We extracted summary-level data for these risk factors from the largest published GWAS including (mostly) participants of European ancestry ^12,13,25–32^ that were published before September 2021. Details for each included data source are presented in Table 1, details about the association models specified by each risk factor GWAS are included in Supplemental Table 1. Due to risk factor transformations that included GWAS performed, estimated MR odds ratios (ORs) correspond to a 1 standard deviation increase in risk factors, expect for age at menarche and age at menopause, for which ORs correspond to a 1-year increase.

**Table 1.**
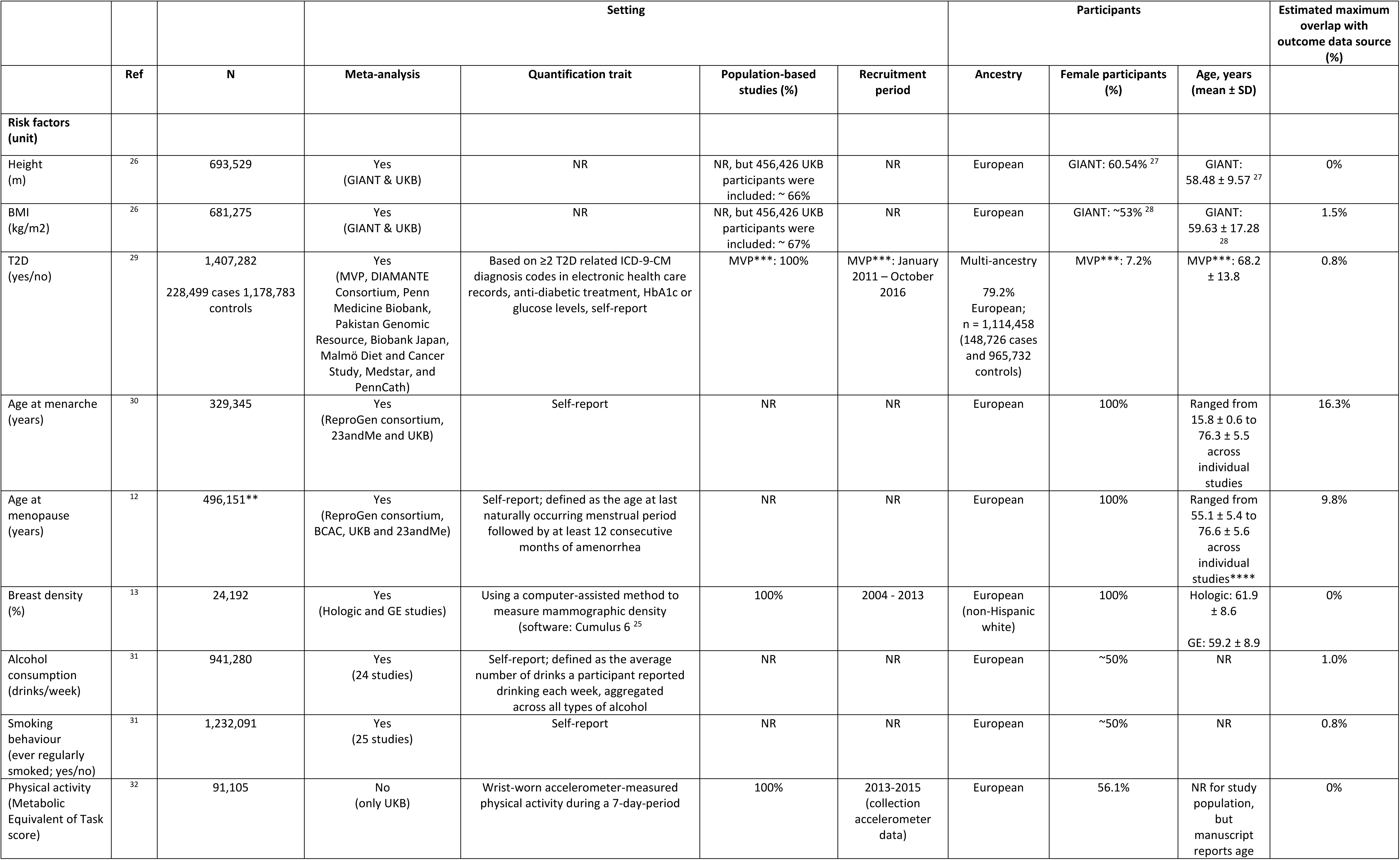
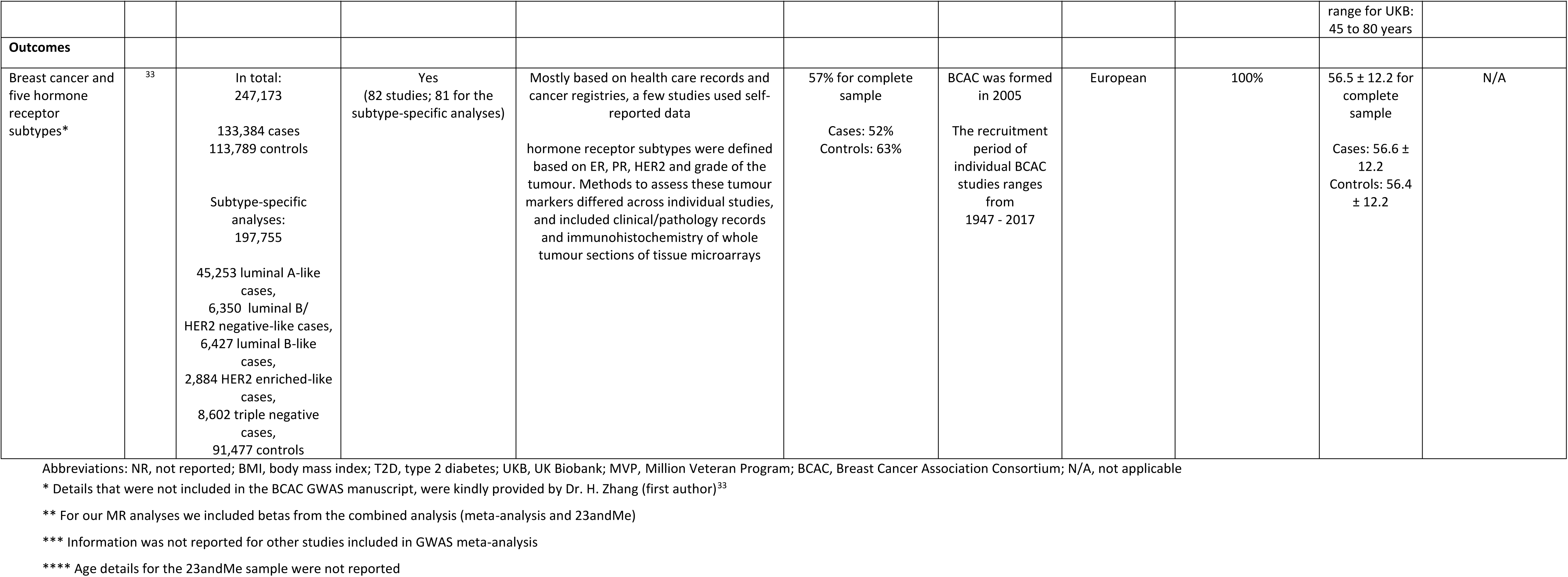
Details about the setting and participants of included risk factor and breast cancer data sources.

### Data source for SNP-breast cancer subtype associations

We extracted publicly available summary-level data from the largest Breast Cancer Association Consortium (BCAC) GWAS to date for total breast cancer (n = 247,173; 133,384 cases, 113,789 controls), and the five hormone receptor breast cancer subtypes: luminal A-like (45,253 cases), luminal B/HER2-negative-like (6,350 cases), luminal B-like (6,427 cases), HER2-enriched-like (2,884 cases), and triple negative breast cancer (8,602 cases)^33^. These subtypes were classified based on tumour grade, ER, progesterone receptor (PR) and human epidermal growth factor receptor 2 (HER2) status as follows: luminal A-like (ER+ and/or PR+ and HER2-, and grade 1 or 2), luminal B/HER2-negative-like (ER+ and/or PR+ and HER2- and grade 3), luminal B-like (ER+ and/or PR+ and HER2+), HER2-enriched-like (ER-,PR-,HER2+), or triple negative (ER-,PR-,HER2-). The same set of controls (91,477 controls) was used for all subtype-specific GWAS analyses. Participants included in this BCAC GWAS were all female and of European ancestry. Additional details about the study population are included in Table 1. SNP-total breast cancer and SNP-subtype associations were estimated using standard logistic regression models and two-stage polytomous logistic models, respectively. Accordingly, the number of cases per hormone receptor subtype represents the effective number of cases per subtype, see Supplementary Note of ^33^ for details. We estimated the maximum percentage of overlap with the selected risk factor GWAS based on the largest number of individuals from studies that were included in both risk factor and BCAC GWAS analyses.

### Selection of risk factor-specific genetic IVs

From each risk factor GWAS, we selected genome-wide significant SNPs (p < 5 x 10^-8^). For height and BMI, an even more stringent p-value threshold (p < 1 x 10^-8^) was used by the GWAS authors for the identification of significant hits ^26^. The T2D GWAS ^29^ was the only data source that included a trans-ethnic population. To avoid confounding due to population stratification ^8^, we only included SNPs that reached genome-wide significance in European ancestry-specific T2D GWAS analyses. For SNPs that were not available in the BCAC GWAS summary statistics, we searched for proxy SNPs (linkage disequilibrium (LD) r^2^ ≥ 0.8) using the NIH LDlink API implemented in the *LDlinkR* R-package ^34^. To maximize statistical power to detect causal effects, we did not exclude SNPs based on pairwise correlations for our primary analyses, but instead accounted for variant correlation in the analysis. Prior to conducting MR analyses, we performed harmonization of alleles and effect estimates between the risk factor and BCAC GWAS using the *TwoSampleMR* R-package ^35^. At this step, we excluded palindromic SNPs with intermediate allele frequencies (i.e., A/T or C/G SNPs with an effect allele frequency ranging from 0.40 to 0.60) because harmonization of these specific variants between different data sources can be error prone. In addition, SNPs that were not available in the 1000G phase 3 reference panel were excluded during harmonization. Supplemental Figure 1 presentsa graphical overview of this selection process.

### Statistical analyses

Prior to performing our primary MR analyses, we set out to calculate LD matrices for all SNPs for the specific risk factors using the *ld_matrix* function implemented in the *TwoSampleMR* R-package. However, due to the substantial proportion of highly correlated SNPs for height and BMI, the correlation matrices that we calculated for these risk factors were near-singular. Therefore, we used a previously published method ^36^ that performs unscaled principal component analysis on a weighted version of the genetic correlation matrix instead. This method results in transformed values for the SNP-risk factor and SNP-outcome associations and a transformed correlation matrix. We included these transformed objects in our primary inverse-variance weighted (IVW) analyses.

#### Primary analysis

We employed the IVW method using a multiplicative random-effects model to calculate primary MR estimates for all nine risk factors in relation to each hormone receptor breast cancer subtype. For these analyses, we included SNP-risk factor estimates from the largest GWAS sample available. Consequently, we used estimates from sex-combined estimates for the risk factors height, BMI, T2D, smoking behaviour, alcohol consumption and physical activity. If analyses from conditional and joint GWAS analyses (i.e., analyses that identify index and independent secondary SNPs) were available, we used these estimates to weigh SNP-risk factor associations (Supplemental Table 1). We performed post-hoc power calculations for subtype-specific associations using a publicly available web application (https://sb452.shinyapps.io/power/). Specifically, we estimated statistical power to detect the magnitude of the causal effect estimate for overall breast cancer, as this estimate would be the most accurate under the null hypothesis that there is no heterogeneity across subtypes (see Supplemental Table 2 for used parameters). In addition to estimating causal effects of each risk factor on total breast cancer and breast cancer subtypes, we calculated the I^2^ index to quantify heterogeneity (%) in primary MR estimates across subtypes. We calculated I^2^ estimates through meta-analysis of the five subtype-specific causal effects estimates per risk factor using random-effect models implemented in the *metafor* R-package ^37^. As our results suggested consistently different effect estimates for triple negative tumours, we calculated heterogeneity in subtype-specific estimates after exclusion of this subtype as post-hoc analysis.

#### Secondary analyses

We additionally performed IVW analyses restricted to uncorrelated SNPs (LD r2 ≤ 0.001). Our rationale for this was two-fold: (1) direct comparison with previously published MR estimates for breast cancer, and (2) direct comparison with robust MR analyses, which are not all extended for the inclusion of correlated IVs.

To yield valid causal estimates, genetic IVs have to meet the relevance, independence and exclusion restriction assumptions. Selection of genetic IVs based on the genome-wide significance threshold is an accepted statistical approach to ensure that SNPs meet the relevance assumption ^38^. To quantify the strength of included genetic IVs, we calculated F-statistics based on r^2^ (i.e., the variance explained in the respective risk factor) estimated in independent study populations, if available (Supplemental Table 3). We performed the following robust MR methods to check how consistent our findings were under less stringent assumptions regarding pleiotropic effects of the included genetic IVs; MR-Egger regression ^39^, weighted median ^40^, mode-based estimator ^41^ and MR-PRESSO^42^. Altogether, the results of these different methods give some insight into the plausibility of the exclusion restriction assumption. Based on previous MR findings regarding breast cancer ^11^, we also performed multivariable MR analyses for BMI and age at menarche. We performed two separate multivariable MR analyses; the first including summary-level data for BMI from the GWAS by Yengo et al. ^26^, and the second including summary-level data from the largest BMI GWAS in UK Biobank by Elsworth et al. ^43^. The reason for this was that ∼25% of the genetic IVs for age at menarche were missing in the most recent BMI GWAS ^26^, whereas all variants were available in the UK Biobank GWAS summary statistics ^43^.

In addition to these three fundamental assumptions, the homogeneity assumption should be reasonable in two-sample MR studies ^23^. However, the BCAC GWAS only included women, whereas six of the nine selected risk factor GWAS included both females and males (Table 1). Accordingly, the homogeneity assumption is not met by design. To assess potential bias because of this violation, we conducted secondary analyses in which we weighed the SNPs that reached genome-wide significance in sex-combined analyses using female-specific betas. For height and BMI, we extracted female-specific betas from a previous GIANT GWAS ^44,45^. Of the 3,146 SNPs for height, only 3,002 SNPs were available in this female-specific data source. For the remaining 144 SNPs we included weights from sex-combined GWAS analyses. For physical activity, we received female-specific betas from the authors of the selected GWAS ^32^. For T2D, alcohol consumption and smoking behaviour female-specific estimates were not publicly available.

We ultimately combined results from our primary and secondary MR analyses to evaluate the strength of evidence for causal effects of each risk factor on each breast cancer subtype. For this evaluation, we used the following recently proposed definition ^46^: evidence was considered to be “robust” if all performed MR methods presented a p-value < 0.05; evidence was considered to be “probable” if at least one method (primary or secondary analysis) had a p-value < 0.05 and the direction of the effect estimate was concordant across all methods; evidence was considered to be “suggestive” if at least one method had a p-value < 0.05, but the direction of the effect estimates differed across methods; and evidence was considered to be “insufficient” if all MR methods had a p-value ≥ 0.05.

All analyses were performed using R version 4.0.5 (https://www.R-project.org/). All R code, including details on package versions, and summary-level data that were used to generate our results are available at https://github.com/SchmidtGroupNKI/MR_BCsubtypes.

## Results

### Descriptive statistics data sources

Details about the setting and participants included in each selected risk factor GWAS and the breast cancer subtype GWAS are presented in Table 1. Total sample size of the included GWAS ranged from 24,192 to 1,232,091 individuals for breast density and smoking behaviour, respectively. Except for the T2D GWAS all data sources were restricted to European study populations (T2D GWAS: 79.2% European). GWAS for height, BMI, T2D, alcohol consumption, regular smoking and physical activity included both females and males. For the latter three risk factors, ∼50% of the study participants was female, for the anthropometric-related risk factors these details were insufficiently reported. The age distribution of study participants included in the breast cancer subtype GWAS and risk factor GWAS were similar with a reported mean age over 55 years. We estimated the maximum overlap in study participants between the risk factor and BCAC GWAS to range from 0% (height, breast density and physical activity) to 16.3% (age at menarche) based on details reported by the included data sources.

### Causal effects of established breast cancer risk factors across hormone receptor breast cancer subtypes

Causal effect estimates (ORs per 1 standard deviation increase or per 1 year increase for age at menarche and age at menopause) for each of the nine breast cancer risk factors across the five hormone receptor breast cancer subtypes are presented in Figure 1 and Table 2. Statistical power estimates corresponding to our primary MR estimates are presented in Supplemental Table 2. In general, we observed that IVW estimates for luminal A-like and luminal B/HER2-negative-like subtypes were very similar to IVW estimates for overall breast cancer. Heterogeneity across subtype-specific causal effects was not due to opposite causal effect estimates, but due to stronger estimates or absence of an effect for some subtypes.

**Figure 1.**
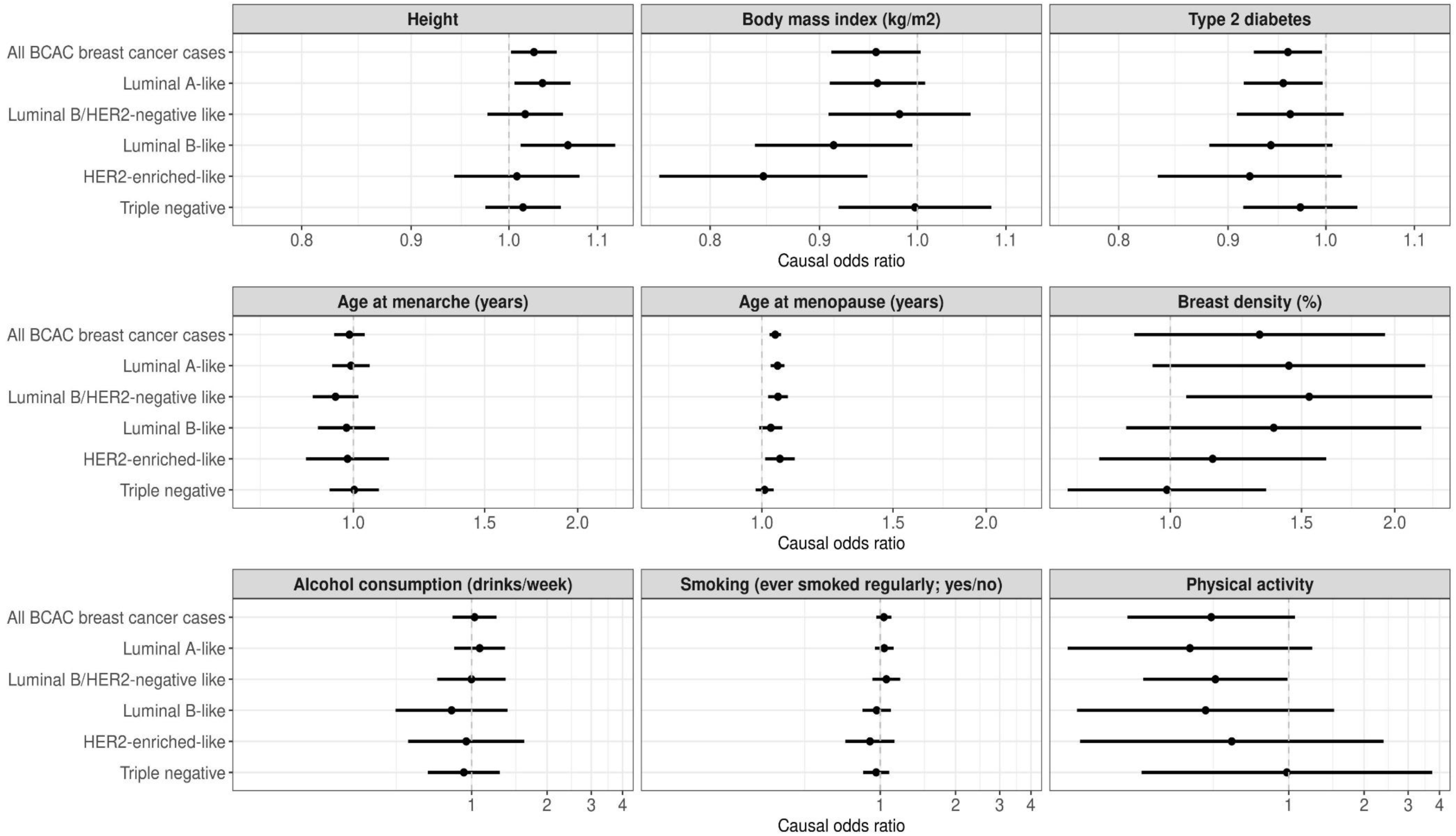
Causal breast cancer subtype-specific effect estimates per unit increase for nine established breast cancer risk factors. Presented ORs and 95% confidence intervals were calculated using the inverse-variance weighted method including correlated variants, which was taken into account through the inclusion of a transformed linkage disequilibrium matrix (see Methods). ORs correspond to a 1 standard deviation increase for all risk factors, except for age at menarche and age at menopause. ORs for these two risk factors correspond to a 1 year increase. The grey vertical dotted line indicates an OR of 1.00 (i.e., absence of a causal association).

**Table 2.** Subtype-specific causal effect estimates per unit* increase for all nine breast cancer risk factors across primary and secondary MR analyses.

| Risk factor | Outcome | Primary IVW analysis |  | Secondary IVW analysis |  | MR-Egger |  |  | Weighted median |  | Weighted mode |  | MR-PRESSO** |  |
| --- | --- | --- | --- | --- | --- | --- | --- | --- | --- | --- | --- | --- | --- | --- |
|  |  | OR | 95%CI | OR | 95%CI | OR | 95%CI | p-value intercept*** | OR | 95%CI | OR | 95%CI | OR | 95%CI |
| **Height** | All BCAC breast cancer cases | **1.03** | **(1.00, 1.05)** | **1.05** | **(1.02, 1.07)** | **1.04** | **(1.00, 1.08)** | 0.69 | 1.02 | (0.99, 1.05) | 1.01 | (0.98, 1.04) | **1.02** | **(1.00, 1.04)** |
|  | Luminal A-like | **1.04** | **(1.01, 1.07)** | **1.07** | **(1.03, 1.10)** | **1.06** | **(1.02, 1.11)** | 0.87 | **1.04** | **(1.00, 1.08)** | **1.04** | **(1.00, 1.08)** | **1.05** | **(1.02, 1.08)** |
|  | Luminal B/HER2-negative like | 1.02 | (0.98, 1.06) | **1.06** | **(1.02, 1.10)** | 1.06 | (1.00, 1.12) | 0.94 | 1.05 | (0.99, 1.12) | 1.04 | (0.98, 1.11) | **1.05** | **(1.01, 1.09)** |
|  | Luminal B-like | **1.07** | **(1.01, 1.12)** | **1.06** | **(1.02, 1.11)** | 1.03 | (0.97, 1.11) | 0.27 | 1.04 | (0.97, 1.13) | 1.01 | (0.93, 1.10) | **1.07** | **(1.02, 1.12)** |
|  | HER2-enriched-like | 1.01 | (0.94, 1.08) | 1.02 | (0.96, 1.09) | 1.05 | (0.96, 1.15) | 0.37 | 0.98 | (0.88, 1.09) | 0.99 | (0.89, 1.10) | NA | NA |
|  | Triple negative | 1.02 | (0.97, 1.06) | 1.03 | (0.99, 1.07) | 1.02 | (0.97, 1.08) | 0.64 | 1.02 | (0.96, 1.09) | 1.00 | (0.94, 1.07) | 1.03 | (0.99, 1.07) |
| **Body mass index (kg/m2)** | All BCAC breast cancer cases | 0.96 | (0.91, 1.00) | **0.95** | **(0.91, 1.00)** | 0.97 | (0.90, 1.05) | 0.54 | 0.96 | (0.91, 1.02) | 0.99 | (0.94, 1.05) | 0.97 | (0.93, 1.00) |
|  | Luminal A-like | 0.96 | (0.91, 1.01) | 0.98 | (0.93, 1.04) | 1.00 | (0.91, 1.10) | 0.60 | 1.00 | (0.93, 1.07) | 1.04 | (0.97, 1.11) | 0.98 | (0.94, 1.02) |
|  | Luminal B/HER2-negative like | 0.98 | (0.91, 1.06) | 0.95 | (0.89, 1.03) | 0.98 | (0.87, 1.11) | 0.61 | 0.99 | (0.88, 1.11) | 0.98 | (0.88, 1.09) | 0.95 | (0.88, 1.02) |
|  | Luminal B-like | **0.91** | **(0.84, 0.99)** | 0.98 | (0.89, 1.07) | 1.04 | (0.89, 1.2) | 0.32 | 1.04 | (0.91, 1.19) | 1.03 | (0.91, 1.17) | 0.97 | (0.89, 1.05) |
|  | HER2-enriched-like | **0.85** | **(0.76, 0.95)** | 0.92 | (0.82, 1.04) | 0.99 | (0.81, 1.21) | 0.37 | 1.03 | (0.84, 1.25) | 1.05 | (0.87, 1.26) | 0.92 | (0.81, 1.03) |
|  | Triple negative | 1.00 | (0.92, 1.08) | 0.98 | (0.91, 1.06) | 1.03 | (0.91, 1.18) | 0.32 | 1.03 | (0.91, 1.17) | 0.98 | (0.87, 1.11) | 0.98 | (0.91, 1.06) |
| **Type 2 diabetes** | All BCAC breast cancer cases | **0.96** | **(0.93, 1.00)** | 1.00 | (0.97, 1.03) | 0.96 | (0.90, 1.02) | 0.14 | 1.02 | (0.99, 1.05) | **1.11** | **(1.06, 1.17)** | 0.99 | (0.97, 1.01) |
|  | Luminal A-like | **0.96** | **(0.92, 1.00)** | 1.00 | (0.96, 1.03) | 0.95 | (0.88, 1.02) | 0.13 | 1.00 | (0.97, 1.04) | **1.13** | **(1.05, 1.22)** | 0.98 | (0.95, 1.01) |
|  | Luminal B/HER2-negative like | 0.96 | (0.91, 1.02) | 1.02 | (0.97, 1.07) | 0.96 | (0.87, 1.05) | 0.13 | 1.05 | (0.98, 1.12) | 1.04 | (0.95, 1.14) | 1.02 | (0.98, 1.07) |
|  | Luminal B-like | 0.94 | (0.88, 1.01) | 0.97 | (0.92, 1.03) | 0.93 | (0.83, 1.03) | 0.31 | 0.99 | (0.90, 1.09) | 0.99 | (0.90, 1.09) | 0.99 | (0.94, 1.04) |
|  | HER2-enriched-like | 0.92 | (0.83, 1.02) | **0.92** | **(0.86, 0.99)** | 0.92 | (0.79, 1.07) | 0.94 | 0.88 | (0.77, 1.01) | 0.90 | (0.76, 1.05) | NA | NA |
|  | Triple negative | 0.97 | (0.91, 1.03) | 1.01 | (0.96, 1.05) | 1.03 | (0.93, 1.13) | 0.65 | 1.07 | (1.00, 1.14) | **1.23** | **(1.11, 1.36)** | 0.99 | (0.95, 1.04) |
| **Age at menarche (years)** | All BCAC breast cancer cases | 0.99 | (0.94, 1.04) | 0.98 | (0.94, 1.02) | 0.98 | (0.88, 1.09) | 0.92 | 0.98 | (0.94, 1.02) | 0.96 | (0.90, 1.03) | 0.97 | (0.94, 1.00) |
|  | Luminal A-like | 0.99 | (0.94, 1.05) | 0.99 | (0.95, 1.04) | 0.98 | (0.86, 1.11) | 0.79 | 0.97 | (0.91, 1.03) | 0.91 | (0.82, 1.01) | 0.98 | (0.94, 1.02) |
|  | Luminal B/HER2-negative like | 0.95 | (0.88, 1.02) | 0.96 | (0.90, 1.03) | 1.02 | (0.86, 1.22) | 0.47 | 0.94 | (0.85, 1.03) | 0.95 | (0.79, 1.14) | 0.95 | (0.90, 1.02) |
|  | Luminal B-like | 0.98 | (0.90, 1.07) | 1.02 | (0.94, 1.10) | 0.94 | (0.77, 1.15) | 0.40 | 1.00 | (0.90, 1.12) | 1.01 | (0.83, 1.22) | 1.00 | (0.93, 1.07) |
|  | HER2-enriched-like | 0.98 | (0.86, 1.12) | 0.97 | (0.87, 1.07) | 0.98 | (0.74, 1.30) | 0.90 | 0.97 | (0.83, 1.14) | 0.89 | (0.68, 1.17) | NA | NA |
|  | Triple negative | 1.00 | (0.93, 1.08) | 0.98 | (0.92, 1.04) | 1.02 | (0.86, 1.21) | 0.58 | 1.00 | (0.90, 1.10) | 1.02 | (0.88, 1.19) | 0.96 | (0.91, 1.02) |
| **Age at menopause (years)** | All BCAC breast cancer cases | **1.04** | **(1.02, 1.06)** | **1.04** | **(1.02, 1.05)** | **1.04** | **(1.00, 1.07)** | 0.91 | **1.05** | **(1.03, 1.06)** | **1.05** | **(1.03, 1.07)** | **1.05** | **(1.04, 1.06)** |
|  | Luminal A-like | **1.05** | **(1.03, 1.07)** | **1.04** | **(1.03, 1.06)** | **1.04** | **(1.00, 1.08)** | 0.82 | **1.06** | **(1.04, 1.08)** | **1.06** | **(1.03, 1.08)** | **1.06** | **(1.04, 1.07)** |
|  | Luminal B/HER2-negative like | **1.05** | **(1.02, 1.08)** | **1.05** | **(1.02, 1.08)** | 1.03 | (0.97, 1.08) | 0.33 | **1.06** | **(1.02, 1.10)** | **1.06** | **(1.01, 1.10)** | **1.06** | **(1.03, 1.08)** |
|  | Luminal B-like | 1.03 | (0.99, 1.07) | **1.03** | **(1.00, 1.06)** | 1.02 | (0.96, 1.08) | 0.68 | 1.04 | (0.99, 1.08) | 1.04 | (0.98, 1.09) | **1.04** | **(1.01, 1.07)** |
|  | HER2-enriched-like | **1.06** | **(1.01, 1.11)** | **1.04** | **(1.00, 1.07)** | 1.05 | (0.98, 1.14) | 0.65 | 1.05 | (0.99, 1.11) | 1.08 | (0.98, 1.18) | NA | NA |
|  | Triple negative | 1.01 | (0.98, 1.04) | 1.00 | (0.98, 1.03) | 1.04 | (0.99, 1.10) | 0.09 | 1.03 | (0.99, 1.07) | 1.04 | (0.99, 1.10) | NA | NA |
| **Breast density (%)** | All BCAC breast cancer cases | 1.32 | (0.89, 1.94) | 1.03 | (0.68, 1.57) | 0.43 | (0.06, 3.17) | 0.38 | 1.18 | (1.00, 1.39) | **1.39** | **(1.16, 1.66)** | **1.26** | **(1.04, 1.51)** |
|  | Luminal A-like | 1.44 | (0.95, 2.20) | 1.09 | (0.68, 1.75) | 0.38 | (0.04, 3.49) | 0.34 | **1.30** | **(1.06, 1.61)** | **1.39** | **(1.06, 1.82)** | 1.19 | (0.89, 1.57) |
|  | Luminal B/HER2-negative like | **1.54** | **(1.05, 2.25)** | 1.05 | (0.71, 1.54) | 0.39 | (0.06, 2.35) | 0.27 | 1.12 | (0.80, 1.56) | 1.40 | (0.65, 3.00) | 1.16 | (0.83, 1.63) |
|  | Luminal B-like | 1.38 | (0.87, 2.17) | 1.07 | (0.68, 1.70) | 0.58 | (0.06, 5.56) | 0.59 | 1.32 | (0.89, 1.95) | 1.93 | (0.94, 3.93) | 1.35 | (0.86, 2.10) |
|  | HER2-enriched-like | 1.14 | (0.80, 1.62) | 0.91 | (0.57, 1.47) | 0.47 | (0.05, 4.81) | 0.56 | 0.93 | (0.56, 1.53) | 0.84 | (0.22, 3.17) | 1.16 | (0.73, 1.82) |
|  | Triple negative | 0.99 | (0.73, 1.34) | 1.01 | (0.63, 1.63) | 0.21 | (0.02, 1.72) | 0.13 | 1.18 | (0.86, 1.63) | 1.39 | (0.87, 2.21) | 1.19 | (0.88, 1.60) |
| **Alcohol consumption (drinks/week)** | All BCAC breast cancer cases | 1.03 | (0.84, 1.26) | 0.92 | (0.77, 1.11) | 0.92 | (0.66, 1.28) | 0.99 | 0.86 | (0.70, 1.05) | 0.88 | (0.73, 1.05) | 0.94 | (0.80, 1.10) |
|  | Luminal A-like | 1.08 | (0.85, 1.36) | 0.91 | (0.73, 1.14) | 0.97 | (0.66, 1.44) | 0.67 | 0.93 | (0.72, 1.21) | 0.92 | (0.73, 1.18) | 0.92 | (0.75, 1.13) |
|  | Luminal B/HER2-negative like | 1.00 | (0.73, 1.37) | 0.90 | (0.62, 1.32) | 0.94 | (0.5, 1.79) | 0.88 | 0.88 | (0.55, 1.40) | 0.86 | (0.56, 1.33) | 0.95 | (0.67, 1.35) |
|  | Luminal B-like | 0.83 | (0.50, 1.39) | 0.78 | (0.54, 1.13) | 0.57 | (0.31, 1.06) | 0.22 | **0.51** | **(0.30, 0.86)** | **0.52** | **(0.32, 0.85)** | 0.75 | (0.54, 1.04) |
|  | HER2-enriched-like | 0.95 | (0.56, 1.62) | 1.35 | (0.84, 2.16) | 1.68 | (0.76, 3.72) | 0.51 | 1.69 | (0.76, 3.74) | 1.64 | (0.77, 3.49) | NA | NA |
|  | Triple negative | 0.93 | (0.67, 1.29) | 0.91 | (0.66, 1.25) | 0.85 | (0.50, 1.45) | 0.76 | 0.81 | (0.51, 1.28) | 0.82 | (0.54, 1.24) | NA | NA |
| **Smoking (ever smoked regularly; yes/no)** | All BCAC breast cancer cases | 1.03 | (0.96, 1.11) | **1.08** | **(1.01, 1.16)** | 1.04 | (0.79, 1.36) | 0.74 | 1.05 | (0.98, 1.13) | 1.00 | (0.83, 1.20) | **1.09** | **(1.03, 1.15)** |
|  | Luminal A-like | 1.04 | (0.95, 1.13) | 1.09 | (1.00, 1.18) | 1.22 | (0.87, 1.71) | 0.48 | 1.03 | (0.94, 1.12) | 0.96 | (0.75, 1.22) | 1.05 | (0.98, 1.13) |
|  | Luminal B/HER2-negative like | 1.06 | (0.93, 1.20) | 1.03 | (0.91, 1.16) | 0.82 | (0.51, 1.33) | 0.35 | 0.97 | (0.83, 1.15) | 0.86 | (0.54, 1.37) | NA | NA |
|  | Luminal B-like | 0.97 | (0.85, 1.10) | 1.10 | (0.97, 1.25) | 1.07 | (0.64, 1.81) | 0.93 | 1.05 | (0.87, 1.27) | 0.94 | (0.58, 1.52) | NA | NA |
|  | HER2-enriched-like | 0.91 | (0.73, 1.14) | 1.13 | (0.93, 1.37) | 0.94 | (0.43, 2.05) | 0.64 | 1.08 | (0.82, 1.42) | 1.03 | (0.48, 2.19) | NA | NA |
|  | Triple negative | 0.96 | (0.86, 1.09) | 1.02 | (0.90, 1.15) | 0.81 | (0.50, 1.33) | 0.36 | 0.98 | (0.83, 1.15) | 1.05 | (0.63, 1.77) | 1.01 | (0.89, 1.13) |
| **Physical activity** | All BCAC breast cancer cases | 0.49 | (0.23, 1.06) | 0.53 | (0.28, 1.00) | 0.03 | (1.23 x 10<sup>-4</sup>, 5.86) | 0.28 | **0.66** | **(0.46, 0.93)** | 0.92 | (0.66, 1.29) | 0.85 | (0.34, 2.05) |
|  | Luminal A-like | 0.40 | (0.13, 1.24) | 0.44 | (0.17, 1.13) | 0.04 | (5.13 x 10<sup>-6</sup>, 321.75) | 0.60 | **0.53** | **(0.33, 0.85)** | 0.82 | (0.49, 1.37) | 0.42 | (0.00, 29.22) |
|  | Luminal B/HER2-negative like | **0.51** | **(0.26, 0.99)** | **0.47** | **(0.24, 0.92)** | 3.65 | (0.01, 1734.32) | 0.51 | 0.66 | (0.34, 1.30) | 0.69 | (0.33, 1.44) | NA | NA |
|  | Luminal B-like | 0.47 | (0.14, 1.52) | 0.49 | (0.19, 1.31) | 4.06 x 10<sup>-3</sup> | (1.34 x 10<sup>-6</sup>, 12.28) | 0.24 | 0.66 | (0.28, 1.59) | 1.21 | (0.28, 5.14) | NA | NA |
|  | HER2-enriched-like | 0.59 | (0.15, 2.39) | 0.61 | (0.20, 1.89) | 1.79 x 10<sup>-3</sup> | (2.07 x 10<sup>-7</sup>, 15.43) | 0.20 | 0.65 | (0.19, 2.24) | 1.26 | (0.18, 9.01) | NA | NA |
|  | Triple negative | 0.98 | (0.26, 3.74) | 0.96 | (0.31, 3.02) | 0.49 | (6.44 x 10<sup>-6</sup>, 36805.73) | 0.91 | 0.77 | (0.37, 1.61) | 0.80 | (0.36, 1.78) | 0.82 | (0.45, 1.49) |

Abbreviations: IVW, inverse-variance weighted; OR, odds ratio; CI, confidence interval; BCAC, breast cancer association consortium; HER2, human epidermal growth factor receptor 2; NA, not applicable Bold figures indicate causal estimates for which p < 0.05
\*ORs correspond to a 1 standard deviation increase for all risk factors, except for age at menarche and age at menopause. ORs for these two risk factors correspond to a 1 year increase.
\*\* ORs for MR-PRESSO method are only presented if the outlier test (p < 0.05) indicated that one or multiple outlying SNPs were distorting the IVW estimate
\*\*\* A p-value < 0.05 for the MR-Egger intercept indicates that unbalanced horizontal pleiotropy is present

#### Anthropometric risk factors

For height, primary IVW estimates were similar across breast cancer subtypes (I^2^=0%), but suggested only a causal risk-increasing effect of increasing height on luminal A-like and luminal B-like tumours. Estimated heterogeneity across hormone receptor subtypes remained 0% after exclusion of the estimate for triple negative tumours. The combination of our primary and secondary MR analyses provided *robust* evidence for a causal risk-increasing effect of increased height on the risk of luminal A-like tumours, and *probable* evidence for causal associations with luminal B/HER2-negative and luminal B-like tumours (Table 2;Supplemental Figure 2A). Evidence for causal associations with HER2-enriched and triple negative tumours was *insufficient*. However, MR analyses including female-specific genetic IVs provided *robust* evidence for a causal risk-increasing effect of increased height on luminal B-like tumours, and *probable* evidence for the other four subtypes (Supplemental Table 4;Figure 2).

**Figure 2.**
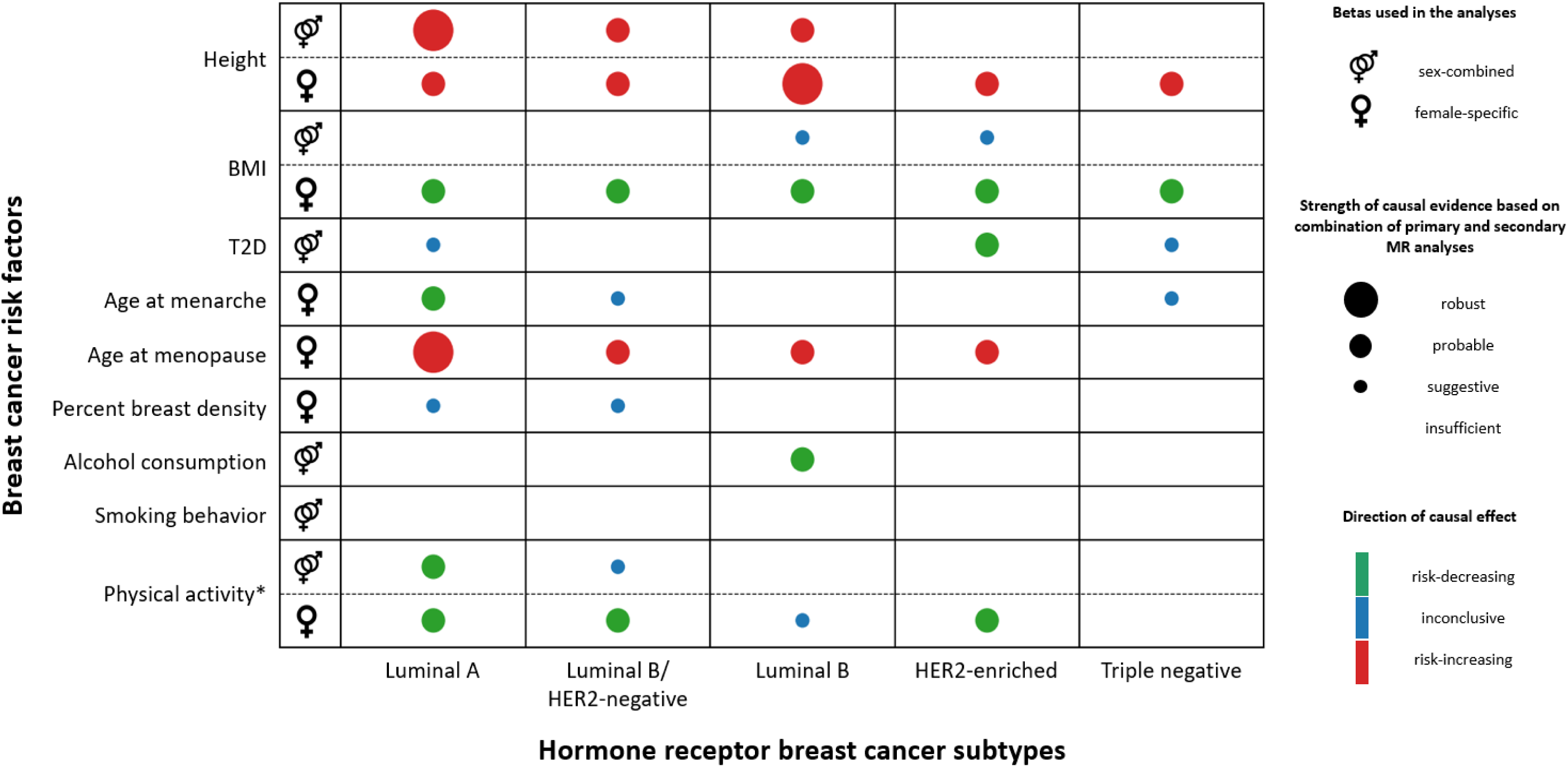
Overview of evidence for (subtype-specific) causal effects per increasing unit of the risk factor. For this figure we counted the number of performed MR methods (both primary and secondary analyses) that provided statistical evidence for causal effects (threshold p < 0.05) and assessed the direction of causal effect estimates. In case the MR-PRESSO analysis indicated that there were no outliers, and thus the secondary IVW estimate was valid, we included the IVW estimate from our secondary analyses twice. Evidence was considered to be *robust* if all performed MR methods for the specific association reached p < 0.05. Evidence was considered to be *probable* if at least one MR method (main or sensitivity) for the specific association reached p < 0.05 and the direction of the effect estimate was concordant for all methods. Evidence was considered to be *suggestive* if at least one MR method (main or sensitivity) for the specific association reached p < 0.05, but the direction of the effect estimate differed between methods. Evidence was considered to be *insufficient* if none of the MR methods reached p < 0.05. * MR-Egger, weighted median, weighted mode and MR-PRESSO analyses are less suitable when only few genetic IVs are available, the results for physical activity should therefore be interpreted with appropriate caution. Abbreviations: BMI, body mass index; T2D, type 2 diabetes; MR, Mendelian randomization

For increasing BMI, primary IVW estimates provided evidence for a causal risk-decreasing effect on luminal B-like and HER2-enriched-like tumours (OR_lumB_=0.91 [95%CI: 0.84, 0.99] and OR_HER2+_=0.85 [95%CI: 0.76, 0.95]). Accordingly, I^2^ estimates indicated moderate heterogeneity across subtype-specific estimates (I^2^=31.1% across all subtypes; I^2^=40.1% after exclusion of triple negative tumours). However, based on the combination of primary and secondary MR analyses, evidence for a causal effect of BMI was merely *suggestive* for luminal B-like and HER2-enriched tumours and insufficient for the other subtypes (Table 2;Supplemental Figure 2B). In contrast, analyses including female-specific genetic IVs provided *probable* evidence for a causal risk-decreasing effect of increasing BMI on all hormone receptor subtypes (Supplemental Table 4;Figure 2).

For increasing risk of T2D, our primary analyses only suggested a causal risk-decreasing effect on luminal A-like tumours, although causal effect estimates for the other subtypes were very similar (I^2^=0% for analyses in- and excluding triple negative tumours). Altogether, our primary and secondary MR analyses provided *probable* evidence for a causal risk-decreasing effect on the risk of HER2-enriched tumours. Evidence for causal associations with luminal A-like and triple negative tumours was *suggestive*, but *insufficient* for luminal B-like and luminal B/HER2-negative-like tumours (Table 2;Supplemental Figure 2C;Figure 2).

#### Reproductive risk factors

We observed no evidence for a causal effect of higher age at menarche on any of the hormone receptor subtypes in primary MR analyses (I^2^=0%). Secondary univariable MR analyses supported these findings (Table 2;Supplemental Figure 2D), but multivariable MR analyses for higher age at menarche and increasing BMI provided evidence for a direct causal association between higher age at menarche and the risk of luminal A-like, luminal B/HER2-negative-like, and triple negative breast tumours (Supplemental Figure 3). However, corresponding heterogeneity estimates suggested similar effects across subtypes (I^2^=15.1% based on Yengo et al. data; 0% based on UK Biobank data). Based on the combination of our primary and secondary analyses, including multivariable MR analyses, evidence for a causal effect of higher age at menarche was only *probable* for luminal A-like tumours, and *suggestive* for luminal B/HER2-negative like and triple negative tumours (Figure 2).

For higher age at menopause, primary IVW estimates suggested causal risk-increasing effects on luminal A-like, luminal B/HER2-negative-like and HER2-enriched, but not on luminal B-like and triple negative tumours (I^2^=42.1%). Estimated heterogeneity across hormone receptor subtypes excluding triple negative breast cancer was 0%, which indicates that the absence of a causal effect on this specific subtype (OR=1.01 [95%CI: 0.98, 1.04]) explains the observed heterogeneity across all five subtypes. Collectively, our MR analyses provided *robust* or *probable* evidence for a causal effect of age at menopause on all subtypes except triple negative breast cancer (Table 2;Supplemental Figure 2E;Figure 2).

In primary IVW analyses, genetically predicted higher breast density was only significantly associated with the risk of luminal B/HER2-negative-like breast cancer (I^2^=15.6% across all subtypes; I^2^=0% after exclusion of triple negative tumours). Based on the combination of all used MR methods, evidence for a causal effect was *suggestive* for luminal A-like and luminal B/HER2-negative like tumours, but *insufficient* for the other subtypes (Table 2;Supplemental Figure 2F;Figure 2).

#### Lifestyle factors

Primary analyses did not provide evidence for causal effects of alcohol consumption and regular smoking on risk of any of the hormone receptor subtypes (I^2^_alcohol_=0%; I^2^_smoking_=0%). Two out of the five secondary MR analyses suggested a causal risk-decreasing effect of higher alcohol consumption on the risk of luminal B-like breast tumours, but not on the other subtypes (Table 2;Supplemental Figure 2G). Consequently, our results only provide *probable* evidence for a causal effect of alcohol consumption of this specific subtype (Figure 2). Secondary analyses for regular smoking supported our primary findings, and thus evidence for a causal association between smoking and risk of any of the hormone receptor subtypes is *insufficient*. For overall higher physical activity, our primary results only provided evidence for a causal risk-decreasing effect on the risk of luminal B-like/HER2-negative breast tumours (I^2^=0%). However, statistical power was in general low, except for the luminal A-like subtype (Supplemental Table 2). Moreover, confidence intervals around primary causal effect estimates were very wide, indicating a high degree of uncertainty. Based on the combination of our primary and secondary MR analyses, we found *probable* evidence for a causal effect of physical activity on luminal A-like breast cancer and *suggestive* evidence for luminal B/HER2-negative-like tumours. For the other subtypes, evidence for a causal effect was *insufficient*. MR analyses including female-specific IVs provided *probable* evidence for a causal risk-decreasing effect on luminal A-like, luminal B/HER2-negative like and HER2-enriched breast tumours (Supplemental Table 4). In these analyses, evidence for a causal association with luminal B-like tumours shifted to *suggestive*, whereas the evidence for a causal association with triple negative tumours remained *insufficient* (Figure 2). Yet, the results of secondary MR analyses for physical activity should be interpreted with caution due to the very low number of genetic IVs available for this risk factor.

## Discussion

This MR study indicates that causal effects of several established breast cancer risk factors differ across hormone receptor breast cancer subtypes. Specifically, we observed moderate heterogeneity in subtype-specific causal effects for age at menopause and breast density. Although this heterogeneity was explained by null findings for triple negative tumours, statistical evidence was also *insufficient* for causal associations of breast density with luminal B-like and HER2-enriched tumours. For height, BMI, risk of T2D, age at menarche, alcohol consumption, regular smoking and physical activity causal effect estimates were similar across breast cancer subtypes. However, only for height and BMI evidence for causal effects was *probable*, or stronger, for all five subtypes. In contrast, for regular smoking statistical evidence for a causal effect was *insufficient* for all subtypes. For the remaining six risk factors, the strength of causal evidence ranged from *probable* to *insufficient* across subtypes. There was no evidence of opposing effects of any risk factor across hormone receptor subtypes. Altogether, our findings suggest that it is more likely that there is heterogeneity in the presence or absence of causal associations between risk factors and breast cancer subtypes, than heterogeneity in the magnitude and direction of causal effects.

Previous MR studies supported height, BMI, age at menarche, age at menopause, breast density and physical activity as breast cancer risk factors, but could not confirm risk of T2D, alcohol consumption and smoking behaviour as causal risk factors for overall breast cancer. The majority of breast cancer patients are diagnosed with luminal tumours ^47^. As a result, established risk factor-breast cancer associations will in general represent associations with luminal subtypes, and potentially less with HER2-enriched and triple negative tumours. Our results indeed indicate that for triple negative breast cancer there is *probable* evidence of causality for only two out of the nine risk factors, while for the other subtypes there is *probable* evidence of causality for four to five risk factors. For HER2- enriched tumours, differences compared with luminal tumours were less clear, possibly due to the considerably lower statistical power for the HER2-enriched subtype. For triple negative tumours, we consider it likely that null findings for age at menopause, breast density and physical activity are not caused by insufficient statistical power, because the sample size for this subtype was similar to that for luminal B/HER2-negative-like and luminal B-like tumours and causal ORs were consistently one.

Considering results from observational studies (i.e., not MR), recent large-scale analyses for height and age at menopause in relation to hormone receptor breast cancer subtypes are in line with our findings. Specifically, increasing height was associated with a higher risk of ER+PR+, ER+PR- and ER-PR- postmenopausal breast cancer ^48^. In a recent BCAC analysis of self-reported data, age at menopause was also not associated with the risk of triple-negative tumours ^1^. Results from the same two studies for BMI and age at menarche were however not in line with our findings. A higher adult BMI was associated with a lower risk of ER+PR+ premenopausal breast cancer and a higher risk of ER+PR+ and ER-PR- postmenopausal breast cancer ^48^. Evidence for associations between BMI and other subtypes was less clear, although an association between higher BMI and a lower risk of breast cancer was suggested for ER+PR- postmenopausal tumours. The results from our analyses including female-specific weights, suggest that a higher genetically-determined BMI is associated with a lower risk of all hormone receptor subtypes. Our finding is in line with an earlier MR study that found that a higher BMI was associated with a lower risk of both pre- and postmenopausal breast cancer ^49^, which contradicts observational evidence showing that a higher BMI is associated with a higher risk of postmenopausal breast cancer (e.g. ^50^). Unfortunately, we were not able to further investigate differences between pre- and postmenopausal status at diagnosis, due to the lack of GWAS summary-level data stratified by menopausal status. Based on observational BCAC data, a younger age at menarche was associated with all hormone receptor subtypes ^1^, whereas our MR results only provided *probable* causal evidence for an association with luminal A-like tumours. Yet, estimated heterogeneity across subtypes in multivariable MR estimates for age at menarche was negligible, and statistical power to detect a causal effect was limited. Two recent observational studies suggested that breast density was also associated with all subtypes ^4,51^, but based on our results evidence for a causal association is weak. Large-scale analyses that studied associations with hormone receptor breast cancer subtypes are currently lacking for risk of T2D, alcohol consumption, smoking behaviour, and physical activity, which hampers a meaningful comparison with our findings for these risk factors. However, systematic reviews of observational studies for several of these lifestyle-related traits indicate that their results for overall breast cancer are likely to be biased by unmeasured confounding(e.g. ^52^). This observation highlights the importance of approaches that are more robust to residual confounding and measurement error, like MR, to understand the aetiology of breast cancer subtypes.

Until now, one other MR analysis set out to investigate multiple known risk factors in relation to hormone receptor breast cancer subtypes ^17^. This previous study reported similar results for age at menopause, which was associated with all subtypes but triple negative tumours. Furthermore, a subtype-specific casual effect for alcohol consumption was reported, which was suggested to be only causally associated with the risk of HER2-enriched breast cancer. However, their MR-PRESSO and MR-Egger estimates for alcohol consumption were in line with our evidence for a causal risk-decreasing effect on the risk of luminal B-like tumours. This contradiction illustrates the added value of our approach to evaluate causal evidence based on the combination of six different MR methods (seven for age at menarche). Consequently, our conclusions are less likely to reflect invalid causal inferences due to unbalanced horizontal pleiotropy or due to chance findings because of multiple testing. We also assessed the homogeneity assumption through the inclusion of female-specific genetic IVs for height, BMI and physical activity and showed that these instruments were consistently associated with stronger causal evidence across all breast cancer subtypes, compared to combined-sex genetic IVs. In line with this observation, our findings for risk of T2D, alcohol consumption and regular smoking should be considered preliminary until female-specific IVs for these risk factors are used in future MR analyses. Altogether, our results underline the importance of an extensive investigation of the MR assumptions.

Another frequently unassessed assumption for two-sample MR analyses is that the risk factor and outcome GWAS samples should be independent, i.e. there should be no overlap in study participants^23^. Since the BCAC includes several studies that also participate in other consortia, this assumption was not completely met in the current analysis. Based on reported details by the included GWAS, we estimated a relatively small overlap in participants ranging from 0% to 16.3%. Bias due to sample overlap in two-sample MR studies arises in analyses that include weak genetic instruments. In the case of minimal sample overlap, this bias will be towards the null and thus rather increase Type 2 error rates than Type 1 error rates ^53^. For each risk factor, we estimated maximum and minimal F-statistics corresponding to primary IVW analyses for luminal A-like and HER2-enriched tumours, respectively. We based these estimations on the variance explained by the included SNPs in an independent study population, if such independent r2 estimates were available. This approach minimized the over-estimation of F-statistics due to winner’s curse bias in the discovery GWAS. Minimum F-statistics for height, BMI, alcohol, and smoking were below the arbitrary threshold of 10, which indicates that weak instrument bias may have biased causal effect estimates for these risk factors towards the null. A last important assumption for two-sample MR studies is that regression models employed for the risk factor GWAS and the outcome GWAS should be adjusted for the same covariates ^23^. Specifically, adjustment for potential confounders in the risk factor GWAS can induce collider bias in two-sample MR analyses. In the current analysis, such bias may have affected our results for breast density, because its GWAS was adjusted for BMI. A higher BMI is associated with decreased breast density ^54^, but without data on the causal association and confounding structure between the genetic IVs, breast density, BMI and breast cancer subtypes it is difficult to evaluate the potential impact on our results ^23,55^.

An additional strength of our study compared to previous MR studies that set out to investigate associations between breast cancer risk factors and the hormone receptor subtypes, is that we maximized statistical power of our primary analyses through the inclusion of as many genetic IVs as possible in combination with LD matrices. Although these correlation matrices were estimated based on genetic data of only ∼400 participants, causal estimates from our primary analyses were very similar to estimates from our secondary, more conservative, analyses. Despite our efforts, our post-hoc power analyses indicated that statistical power to detect causal risk factors across all subtypes was still suboptimal. Future MR studies including stronger genetic IVs, i.e., genetic IVs explaining more variance in the risk factor of interest, could further increase statistical power. However, identification of additional loci requires even larger GWAS study populations, and this has proved to be challenging for lifestyle-related risk factors like alcohol consumption, smoking and physical activity. Another possibility to increase statistical power is the inclusion of larger numbers of breast cancer cases for hormone receptor subtypes, which will be possible through initiatives like the Confluence project ^56^. Further improvement of the quality of MR studies can be achieved if future risk factor GWAS stratify analyses by biological sex, and report full details about the included GWAS setting, participants and methods.

In conclusion, our results suggest that, of the established breast cancer risk factors, height and BMI are likely to exert similar causal effects across all breast cancer subtypes. Our MR analyses also suggest that the majority of established breast cancer risk factors are not causally associated with the risk of triple negative tumours. These insights are valuable for the development of primary prevention strategies, and the improvement of breast cancer risk stratification in the general population. Our findings also emphasize the importance of taking breast cancer subtype into account for the identification of novel breast cancer risk factors.

## Data Availability

All R code, including details on package versions, and summary-level data that were used to generate our results are available at https://github.com/SchmidtGroupNKI/MR_BCsubtypes

## Acknowledgements

The authors thank dr. Haoyu Zhang for providing additional details about the BCAC GWAS summary statistics, and professor Aiden Doherty for providing female-specific summary statistics for overall physical activity. We thank Renee Menezes for help with MR-PRESSO. We acknowledge the Breast Cancer Association Consortium (BCAC) for providing summary data.

## Additional information

### Authors’ contributions

Conceived the work that led to submission: RMGV, SB, MKS; Performed analyses: RMGV, MS;

Played an important role in interpreting the results: all authors; Drafted manuscript: RMGV, MS;

Revise manuscript: all authors;

Approved final version of the manuscript: all authors;

Agreed to be accountable for all aspects of the work in ensuring that questions related to the accuracy or integrity of any part of the work are appropriately investigated and resolved; all authors

### Ethics approval and consent to participate

All GWAS that generated the used summary-level data received ethical approval from qualified institutional boards and all study participants provided informed consent. See corresponding manuscripts for original statements.

### Consent for publication

Not applicable; this manuscript does not contain individual-level data.

### Data availability

The code supporting the results reported in this manuscript will be made available upon publication at: https://github.com/SchmidtGroupNKI/MR_BCsubtypes.

## List of abbreviations

BCAC: breast cancer association consortium
BMI: body mass index
ER: estrogen receptor
GIANT: Genetic Investigation of ANthropometric Traits
GWAS: genome-wide association studies
HER2: human epidermal growth factor receptor 2
IV: instrumental variable
IVW: inverse-variance weighted
LD: linkage disequilibrium
MR: Mendelian randomization
PR: progesterone receptor
SNP: single nucleotide polymorphism
T2D: type 2 diabetes

